# The evaluation of a newly developed antigen test (QuickNavi™-COVID19 Ag) for SARS-CoV-2: A prospective observational study in Japan

**DOI:** 10.1101/2020.12.27.20248876

**Authors:** Yuto Takeuchi, Yusaku Akashi, Daisuke Kato, Miwa Kuwahara, Shino Muramatsu, Atsuo Ueda, Shigeyuki Notake, Koji Nakamura, Hiroichi Ishikawa, Hiromichi Suzuki

**Affiliations:** Department of Infectious Diseases, University of Tsukuba Hospital, 2-1-1 Amakubo, Tsukuba, Ibaraki 305-8576, Japan; Division of Infectious Diseases, Department of Medicine, Tsukuba Medical Center Hospital, 1-3-1 Amakubo Tsukuba, Ibaraki 305-8558, Japan; Akashi Internal Medicine Clinic, 3-1-63 Asahigaoka, Kashiwara, Osaka 582-0026, Japan; Denka Co., Ltd. Gosen site, Reaserch & Development Division, Reagent R&D Depertment, 1-2-2 Minami-hon-cho, Gosen-shi, Niigata 959-1695, Japan; Department of Clinical Laboratory, Tsukuba Medical Center Hospital, 1-3-1 Amakubo, Tsukuba, Ibaraki 305-8558, Japan; Department of Respiratory Medicine, Tsukuba Medical Center Hospital, 1-3-1 Amakubo, Tsukuba, Ibaraki 305-8558, Japan; Department of Infectious Diseases, Faculty of Medicine, University of Tsukuba, 1-1-1 Tennodai, Tsukuba, Ibaraki 305-8575, Japan

**Keywords:** antigen test, COVID-19, immunochromatography, QuickNavi™-COVID19 Ag, SARS-CoV-2

## Abstract

**Introduction:** Several antigen tests for severe acute respiratory syndrome coronavirus 2 (SARS-CoV-2) have been developed worldwide, but their clinical utility has not been well established. In this study, we evaluated the analytical and clinical performance of QuickNavi™-COVID19 Ag, a newly developed antigen test in Japan.

**Methods:** This prospective observational study was conducted at a PCR center between October 7 and December 5, 2020. The included patients were referred from a local public health center and 89 primary care facilities. We simultaneously obtained two nasopharyngeal samples with flocked swabs; one was used for the antigen test and the other for real-time reverse transcription PCR (RT-PCR). Using the results of real-time RT-PCR as a reference, the performance of the antigen test was evaluated.

**Results:** A total of 1186 patients were included in this study, and the real-time RT-PCR detected SARS-CoV-2 in 105 (8.9%). Of these 105 patients, 33 (31.4%) were asymptomatic. The antigen test provided a 98.8% (95% confident interval [CI]: 98.0%-99.4%) concordance rate with real-time RT-PCR, along with a sensitivity of 86.7% (95% CI: 78.6%-92.5%) and a specificity of 100% (95% CI: 99.7%-100%). False-negatives were observed in 14 patients, 8 of whom were asymptomatic and had a low viral load (cycle threshold (Ct) >30). In symptomatic patients, the sensitivity was 91.7% (95% CI: 82.7%-96.9%).

**Conclusion:** QuickNavi™-COVID19 Ag showed high specificity and sufficient sensitivity for the detection of SARS-CoV-2. This test is a promising potential diagnostic modality especially in symptomatic patients.

## Introduction

The pandemic of severe acute respiratory syndrome coronavirus 2 (SARS-CoV-2), which causes coronavirus disease 2019 (COVID-19), has laid a detrimental burden on the healthcare system [1]. The effective isolation and early treatment of SARS-CoV-2 patients require rapid and accurate diagnostic methods [2].

Nucleic acid amplification tests (NAATs) for upper respiratory samples have been the mainstay for the identification of infected individuals [3]. However, while these assays are considered the gold-standard examinations, the disadvantages of their finite availability, long turnaround time, and need for skilled technicians have limited their clinical utility [4]. The number of patients eligible to undergo these tests may overwhelm the test capacity in outbreak settings [3]. Antigen tests are cheaper, more accessible point-of-care tests and take a shorter time to produce results; they can therefore be more useful in limited-resource settings, provided they reliably detect SARS-CoV-2.

The reported sensitivity of antigen tests has ranged from 0%-94%, whereas the specificity is consistently high at >97% [3]. QuickNavi™-COVID19 Ag (Denka Co., Ltd., Tokyo, Japan) is a newly developed antigen test in Japan and employs a sandwich immunochromatography method with mouse monoclonal antibodies against SARS-CoV-2. The test result is available within 15 minutes after samples diluted in the buffer have been placed in a well of the test kit. Nevertheless, no study has yet examined its utility.

In the present study, we evaluated the analytical and clinical performance of QuickNavi™-COVID19 Ag using prospectively collected clinical samples. Furthermore, we analyzed the factors that might influence the sensitivity and specificity.

## Patients and methods

We prospectively performed the study between October 7 and December 5, 2020, at a PCR center in Tsukuba, located in the southern part of Ibaraki Prefecture, Japan. During the COVID-19 endemic period, sample-collecting for PCR in the Tsukuba district was intensively performed with a drive-through-type method at the PCR center in Tsukuba Medical Center Hospital (TMCH). During the study period, additional samples for antigen test were collected from patients who have been referred from a local public health center and 89 primary care facilities (Supplementary Table 1) and healthcare workers of TMCH, and their clinical information was obtained after receiving the subjects’ informed consent. If patients had no clinical data, we excluded them from this study. In cases where patients participated in the current study more than once, only the first evaluation was included in this study.

The ethics committee of TMCH approved the present study (approval number: 2020-033).

### Sample collection and procedures for antigen test

For sample collections, we simultaneously obtained two nasopharyngeal samples for antigen test and PCR examination with FLOQSwab™ (Copan Italia S.p.A., Brescia, Italy) as previously described [5]. Antigen test was performed immediately after sample collection according to the manufacturers’ instructions, described in Supplementary Figure 1, and the results were obtained by the visual interpretation of each examiner. Another swab sample was diluted in 3 mL of Universal Transport Medium™ (UTM™) (Copan Italia S.p.A., Brescia, Italy), and the UTM™ was transferred to an in-house microbiology laboratory located next to the drive-through sample-collecting place of the PCR center within an hour of sample collection.

### PCR examinations for SARS-CoV-2 in this study

After the arrival of the UTM™ samples, purification and RNA extraction were performed with magLEAD 6gC (Precision System Science Co., Ltd., Chiba, Japan) from 200 µL aliquots of UTM™ for in-house reverse transcription PCR (RT-PCR) on the same day as sample collection. The RNA was eluted in 100 µL and stored at -80 °C after in-house RT-PCR. The eluted samples were transferred to Denka Co., Ltd., every week for reference real-time RT-PCR of SARS-CoV-2 using a method developed by the National Institute of Infectious Diseases, Japan [6]. If discordance was recognized between the reference real-time RT-PCR and in-house RT-PCR, a re-evaluation was performed with a BioFire^®^ Respiratory Panel 2.1 and FilmArray^®^ systems (BioFire Diagnostics, LLC, UT, USA), and the final judgment was made.

### Limit of detection of QuickNavi™-COVID19 Ag

The limit of detection of QuickNavi™-COVID19 Ag was investigated as follows: the 2019-nCoV/JPN/TY/WK-521 strain (4.2×10^5^ TCID_50_/mL) cultured in VeroE6/TMPRSS2 cells were diluted two-fold stepwise with QuickNavi™ specimen buffer and used as samples. Each sample with different concentrations was tested in triplicate. As shown in Table 1, the limit of detection was 5.3×10^1^ TCID_50_/mL and was consistent throughout the test.

**Table 1.** The limit of detection test results of three repetitive tests for each sample

| Concentration | Results |  |  |
| --- | --- | --- | --- |
| (TCID<sub>50</sub>/mL) | Sample A | Sample B | Sample C |
| 2.1×10<sup>2</sup> | + | + | + |
| 1.1×10<sup>2</sup> | + | + | + |
| 5.3×10<sup>1</sup> | + | + | + |
| 2.6×10<sup>1</sup> | - | - | - |

TCID<sub>50</sub>, median tissue culture infectious dose

### Statistical analyses

The sensitivity and specificity of antigen test were calculated using the Clopper and Pearson method, with 95% confident intervals (CIs). Categorical variables were compared by Fisher’s exact test. P-values <0.05 were considered to represent statistically significant differences. All calculations were conducted using the R 3.3.1 software program (The R Foundation, Vienna, Austria).

## Results

Of the 2079 referred patients and 45 healthcare workers, a total of 1208 individuals who had nasopharyngeal samples collected for antigen test and had provided their informed consent were initially included. We excluded the patients who were duplicates (n=18) or missing symptom data (n=4). We finally included 1186 subjects for the analysis.

Most samples were obtained at the drive-through PCR center, and only 15 were obtained after hospitalization. Of the 1186 subjects, SARS-CoV-2 was detected in 105 (8.9%) by reference real-time RT-PCR. There was one discordant sample that showed positivity on in-house RT-PCR and negativity on reference real-time RT-PCR. The sample was deemed negative by an additional BioFire^®^ Respiratory Panel 2.1 examination. Of the 105 subjects, 72 (68.6%) were symptomatic, and 33 (31.4%) were asymptomatic (Table 2a).

**Table 2a.** Demographic data of the whole study population and cases infected with SARS-CoV-2

|  | Total | SARS-CoV-2 |  |
| --- | --- | --- | --- |
|  |  | Positive | Negative |
| n | 1186 | 105 | 1081 |
| Age (years, median [IQR]) | 36.5 [23.0, 50.0] | 47.0 [30.0, 58.0] | 36.0 [23.0, 49.0] |
| <18 | 164 (13.8) | 11 (10.5) | 153 (14.2) |
| 18-64 | 898 (75.7) | 79 (75.2) | 819 (75.8) |
| ≥ 65 | 124 (10.5) | 15 (14.3) | 109 (10.1) |
| Sex (Female, %) | 539 (45.4) | 43 (41.0) | 496 (45.9) |
| Asymptomatic patients | 415 (35.0) | 33 (31.4) | 382 (35.3) |
| Symptomatic patients | 771 (65.0) | 72 (68.6) | 699 (64.7) |

SARS-CoV-2, severe acute respiratory syndrome coronavirus 2

The characteristics of the symptomatic subjects and cases infected with SARS-CoV-2 are described in Table 2b. Of the symptomatic SARS-CoV-2-positive cases (n=72), the most common symptom was fever (72.2%), followed by cough or sputum production (41.7%), sore throat (23.6%), fatigue (18.1%) and headache (18.1%).

**Table 2b.**
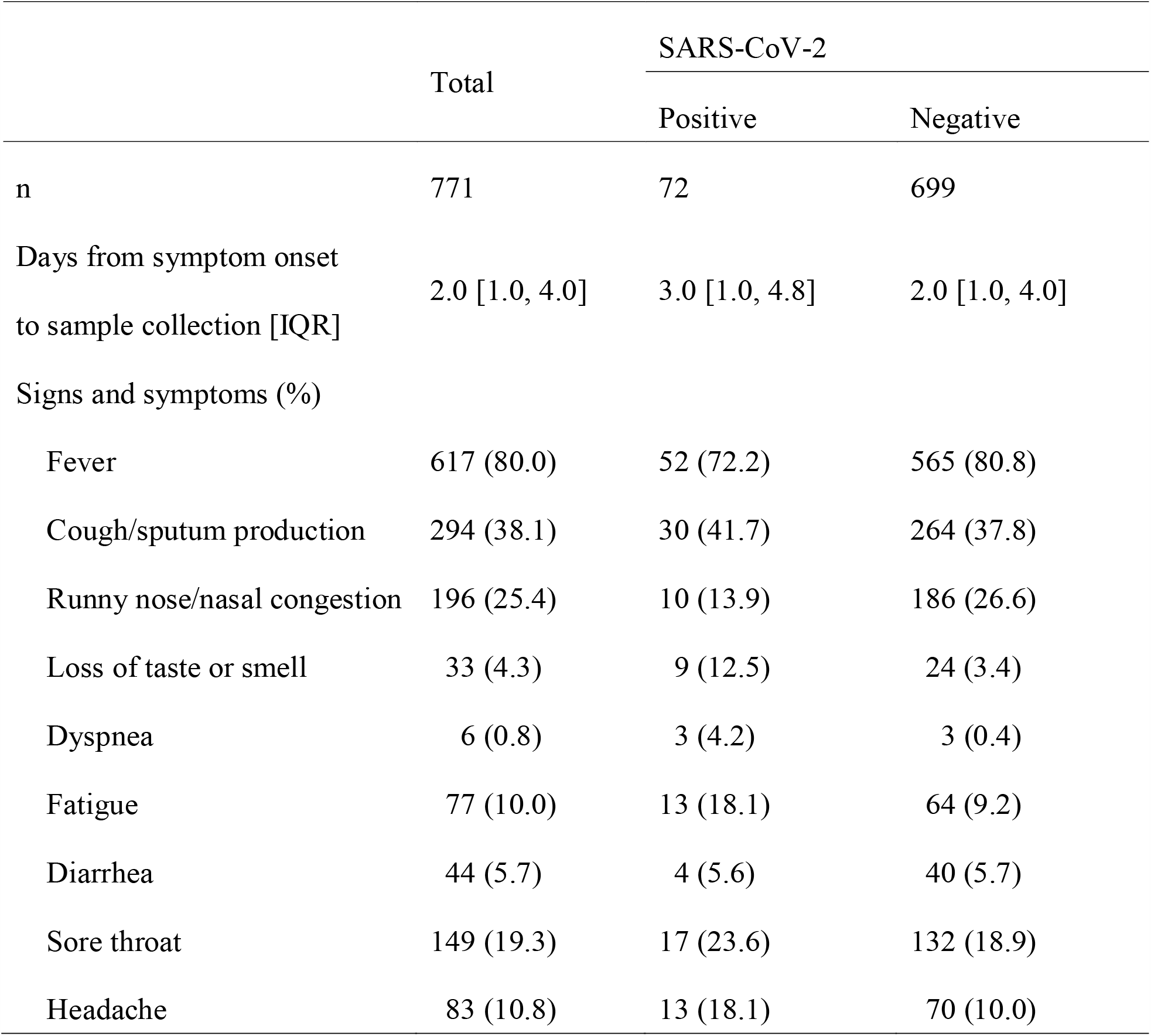
Characteristics of symptomatic patients and cases infected with SARS-CoV-2

### Sensitivity and specificity of antigen test

Of the 105 cases that were positive on reference real-time RT-PCR, antigen test was also positive in 91 (Table 3a). The concordance rate between antigen test and real-time RT-PCR was thus 98.8% (95% CI: 98.0%-99.4%). The sensitivity and specificity rates were 86.7% (95% CI: 78.6%-92.5%) and 100% (95% CI: 99.7%-100%), respectively (Table 3a).

**Table 3a.** Sensitivity and specificity of the QuickNavi™-COVID19 Ag among overall patients

|  |  | real-time RT-PCR |  |
| --- | --- | --- | --- |
|  |  | Positive | Negative |
| Antigen test | Positive | 91 | 0 |
|  | Negative | 14 | 1081 |
| Sensitivity (%) |  | 86.7 (78.6-92.5) |  |
| Specificity (%) |  | 100 (99.7-100) |  |
| Positive predictive value (%) |  | 100 (96.0-100) |  |
| Negative predictive value (%) |  | 98.7 (97.9-99.3) |  |

Of the 72 symptomatic cases that were positive on reference real-time RT-PCR, antigen test was also positive in 66 (Table 3b). The sensitivity and specificity were 91.7% (95% CI: 82.7%-96.9%) and 100% (95% CI: 99.5%-100%), respectively (Table 3b).

**Table 3b.** Sensitivity and specificity of the QuickNavi™-COVID19 Ag among symptomatic patients

|  |  | real-time RT-PCR |  |
| --- | --- | --- | --- |
|  |  | Positive | Negative |
| Antigen test | Positive | 66 | 0 |
|  | Negative | 6 | 699 |
| Sensitivity (%) |  | 91.7 (82.7-96.9) |  |
| Specificity (%) |  | 100 (99.5-100) |  |
| Positive predictive value (%) |  | 100 (94.6-100) |  |
| Negative predictive value (%) |  | 99.1 (98.2-99.7) |  |

Sensitivity, specificity, positive predictive value, and negative predictive value are provided with 95% confident intervals.
RT-PCR, reverse transcription polymerase chain reaction

### Detailed data of discrepant cases between antigen test and real-time RT-PCR examinations

Among the 14 discrepant cases, 8 were asymptomatic, and 4 of the 6 symptomatic cases had their nasopharyngeal samples taken ≥6 days after the onset of symptoms. The N2-gene was detected in all cases, but the N1-gene was not detected in 7 cases. One patient had a history of preceding favipiravir administration (Table 4).

**Table 4.** Detailed data of the 14 cases with discrepant findings between antigen test and real-time RT-PCR

| Case number | Symptoms | Days from the symptom onset to sample collection | Ct value |  | Notes |
| --- | --- | --- | --- | --- | --- |
|  |  |  | N1* | N2* |  |
| 1 | + | 7 | ND | 34 |  |
| 2 | + | 3 | ND | 40 |  |
| 3 | + | 6 | 31 | 24 |  |
| 4 | + | 6 | 35 | 30 | Preceding favipiravir administration |
| 5 | + | 4 | 38 | 35 |  |
| 6 | - | NA | ND | 36 |  |
| 7 | - | NA | ND | 39 |  |
| 8 | - | NA | ND | 37 |  |
| 9 | - | NA | 37 | 31 |  |
| 10 | - | NA | ND | 38 |  |
| 11 | + | 7 | 36 | 30 |  |
| 12 | - | NA | 35 | 30 |  |
| 13 | - | NA | 41 | 39 |  |
| 14 | - | NA | ND | 34 |  |

\*Real-time reverse transcription polymerase chain reaction examinations of SARS-CoV-2 developed by the National Institute of Infectious Diseases, Japan [6].
Ct, cycle threshold; NA, not available; ND, not detected

### Change of sensitivities of antigen test stratified by cycle threshold (Ct) value

The sensitivity of Ct value (N2) <20 was 100% (95% CI: 91.0%-100%), that of Ct 20-24 was 96.7% (95% CI: 82.8%-99.9%), and that of Ct 25-29 was 100% (95% CI: 83.2%-100%) (Table 5). In contrast, the sensitivity of Ct ≥30 was 18.8% (95% CI: 4.0%-45.6%) (Table 5).

**Table 5.**
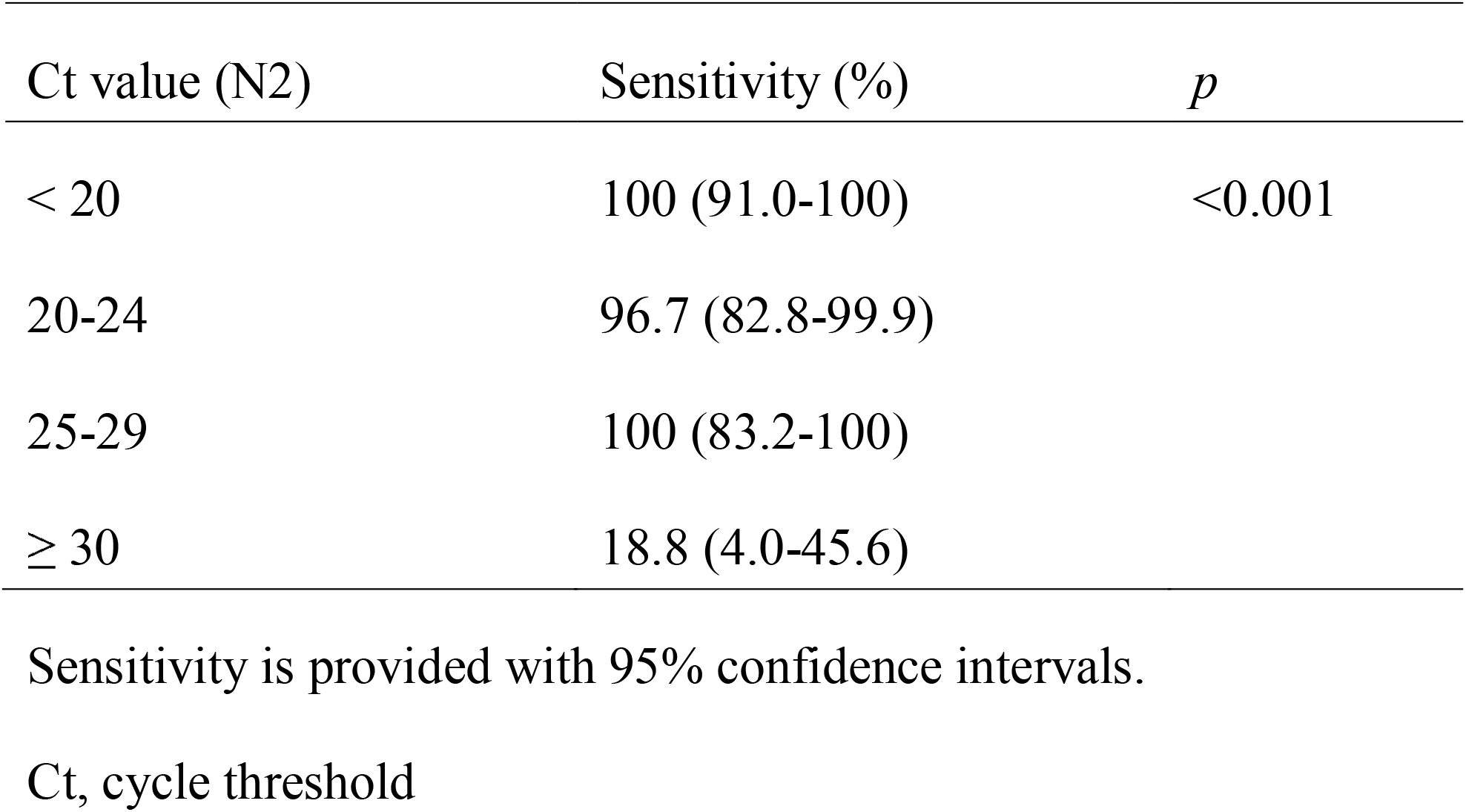
Sensitivities of antigen test stratified by Ct value

## Discussion

Among 1186 subjects referred from clinics and a local healthcare center in the southern part of Ibaraki Prefecture, Japan, this prospective study indicated that QuickNavi™-COVID19 Ag has satisfactory performance for the detection of SARS-CoV-2. Of note, the test provided no false-positive results in our study population. False-negatives were detected in 14 subjects, over half of whom were asymptomatic.

False-positives should be avoided due to concerns about unnecessary further examinations or application of quarantine measures [7]. NAATs are highly specific for SARS-CoV-2, and positive results are usually definitive for the diagnosis of COVID-19 [3]. False-positives are rare and they tend to only be observed under exceptional conditions such as cross contaminations, erroneous handling of samples, or a breakdown in test reagents or equipment [8]. Similar to NAATs, antigen tests generally have high specificities of >99% [9]. Nevertheless, some false-positive results have been reported in other antigen tests [10,11]. While definitive proof is lacking, possible causes for the false-positives include the high viscosity of specimens and interference of human antibodies [12]. The QuickNavi™-COVID19 Ag provided 100% specificity in our study, which exceeded the performance recommended by the World Health Organization (WHO) [3]. Still, whether or not a similar result can be obtained in different settings needs to be confirmed.

The sensitivity of QuickNavi™-COVID19 Ag was 86.7% overall, and the positive detection rate was comparable to the real-time RT-PCR in patients with Ct <30. Eight of 14 false-negative subjects had no symptoms and a low viral load, although conflicting evidence exists regarding the relationship between symptom severity and viral shedding [13,14]. All samples were collected from a nasopharynx with flocked swabs, which may have increased the viral load and improved antigen test sensitivity in our study. The viral load on the nasopharynx is generally higher than in the nasal cavity or saliva [15,16], and flocked swabs can yield more samples than rayon swabs [17].

The utility of antigen tests for screening purposes is controversial. The WHO guidelines basically recommend against antigen tests for screening purposes [3]. In contrast, European countries allow antigen tests for screening or serial testing [18]. Recent studies may support this use of antigen tests, showing the frequency and turnaround time of the tests to be great contributors to an effective screening strategy [19]. Since the QuickNavi™-COVID19 Ag may effectively identify highly infectious patients (generally Ct <25 [20]) without any false-positives, the test may be beneficial for screening purposes.

Several limitations associated with the present study warrant mention. First, reference real-time RT-PCR examinations employed frozen samples. Despite all samples being frozen at -80 °C, their viral load may have been reduced through the storage process. Second, although we investigated whether or not the intervals between the symptom onset and examination timing influenced the performance of the antigen test, the sample size was not sufficient to draw a definitive conclusion (Supplementary Figure 2). Third, using anterior nasal samples was beyond the scope of this study. Sample collection from the anterior nasal cavity is less invasive than that from the nasopharynx and is now approved for QuickNavi™-COVID19 Ag [21]. The clinical performance of the test with these samples has not yet been evaluated, and further research is necessary.

In conclusion, the QuickNavi™-COVID19 Ag showed very high specificity and sufficient sensitivity for the detection of SARS-CoV-2. Given the simple procedures and shorter turnaround time involved with this test, it is a promising option as an alternative diagnostic modality especially in symptomatic patients.

## Data Availability

The data are not publicly available due to their containing information that could compromise the privacy of research participants.

## Acknowledgments

We thank Mrs. Yoko Ueda, Mrs. Mio Matsumoto, and the staff in the Department of Clinical Laboratory of Tsukuba Medical Center Hospital for their intensive support of this study. Mrs. Yoko Ueda and Mrs. Mio Matsumoto significantly contributed to creating the database of this study.

## Figure legends

**Supplementary Figure 1**. Manufacturers’ instructions of QuickNavi™-COVID19 Ag

**Supplementary Figure 2**. Difference in sensitivity of the antigen testing stratified by the day after symptoms onset

## References

[1] World Health Organization. Coronavirus Disease (COVID-19) Situation Reports, https://www.who.int/emergencies/diseases/novel-coronavirus-2019/situation-reports [accessed 22 December 2020].

[2] World Health Organization. Laboratory testing for coronavirus disease (COVID-19) in suspected human cases: interim guidance, 19 March 2020, https://apps.who.int/iris/handle/10665/331501 [accessed 22 December 2020].

[3] World Health Organization. Antigen-detection in the diagnosis of SARS-CoV-2 infection using rapid immunoassays, https://www.who.int/publications/i/item/antigen-detection-in-the-diagnosis-of-sars-cov-2infection-using-rapid-immunoassays [accessed 12 December 2020].

[4] Younes N, Al-Sadeq DW, AL-Jighefee H, Younes S, Al-Jamal O, Daas HI, et al. Challenges in laboratory diagnosis of the novel coronavirus SARS-CoV-2. Viruses 2020;12:582. https://doi.org/10.3390/v12060582.

[5] Marty FM, Chen K, Verrill KA. How to obtain a nasopharyngeal swab specimen. N Engl J Med 2020;382:e76. https://doi.org/10.1056/NEJMvcm2010260.

[6] Shirato K, Nao N, Katano H, Takayama I, Saito S, Kato F, et al. Development of genetic diagnostic methods for detection for novel coronavirus 2019 (nCoV-2019) in Japan. Jpn J Infect Dis 2020;73:304–7. https://doi.org/10.7883/yoken.JJID.2020.061.

[7] Ogawa T, Fukumori T, Nishihara Y, Sekine T, Okuda N, Nishimura T, et al. Another false-positive problem for a SARS-CoV-2 antigen test in Japan. J Clin Virol 2020;131:104612. https://doi.org/10.1016/j.jcv.2020.104612.

[8] World Health Organization. Diagnostic testing for SARS-CoV-2, https://www.who.int/publications/i/item/diagnostic-testing-for-sars-cov-2 [accessed 22 December 2020].

[9] Dinnes J, Deeks JJ, Adriano A, Berhane S, Davenport C, Dittrich S, et al. Rapid, point-of-care antigen and molecular-based tests for diagnosis of SARS-CoV-2 infection. Cochrane Database Syst Rev 2020;8:CD013705. https://doi.org/10.1002/14651858.CD013705.

[10] Tanimoto T, Matsumura M, Tada S, Fujita S, Ueno S, Hamai K, et al. Need for a high-specificity test for confirming weakly positive result in an immunochromatographic SARS-CoV-2-specific antigen test: A case report. J Microbiol Immunol Infect S1684-1182(20)30272-3. https://doi.org/10.1016/j.jmii.2020.11.004.

[11] Chaimayo C, Kaewnaphan B, Tanlieng N, Athipanyasilp N, Sirijatuphat R, Chayakulkeeree M, et al. Rapid SARS-CoV-2 antigen detection assay in comparison with real-time RT-PCR assay for laboratory diagnosis of COVID-19 in Thailand. Virol J 2020;17:177. https://doi.org/10.1186/s12985-020-01452-5.

[12] U.S. Food and Drug Administration. Potential for false positive results with antigen tests for rapid detection of SARS-CoV-2 - Letter to clinical laboratory staff and health care providers, https://www.fda.gov/medical-devices/letters-health-care-providers/potential-false-positive-results-antigen-tests-rapid-detection-sars-cov-2-letter-clinical-laboratory [accessed 22 December 2020].

[13] Magleby R, Westblade LF, Trzebucki A, Simon MS, Rajan M, Park J, et al. Impact of SARS-CoV-2 viral load on risk of intubation and mortality among hospitalized patients with coronavirus disease 2019. Clin Infect Dis 2020;ciaa851. https://doi.org/10.1093/cid/ciaa851.

[14] Lee S, Kim T, Lee E, Lee C, Kim H, Rhee H, et al. Clinical course and molecular viral shedding among asymptomatic and symptomatic patients with SARS-CoV-2 infection in a community treatment center in the Republic of Korea. JAMA Intern Med2020;180:1447. https://doi.org/10.1001/jamainternmed.2020.3862.

[15] Pinninti S, Trieu C, Pati SK, Latting M, Cooper J, Seleme MC, et al. Comparing nasopharyngeal and midturbinate nasal swab testing for the identification of severe acute respiratory syndrome coronavirus 2. Clin Infect Dis 2020;ciaa882. https://doi.org/10.1093/cid/ciaa882.

[16] Procop GW, Shrestha NK, Vogel S, Sickle KV, Harrington S, Rhoads DD, et al. A direct comparison of enhanced saliva to nasopharyngeal swab for the detection of SARS-CoV-2 in symptomatic patients. J Clin Microbiol 2020;58:e01946–20. https://doi.org/10.1128/JCM.01946-20.

[17] Daley P, Castriciano S, Chernesky M, Smieja M. Comparison of flocked and rayon swabs for collection of respiratory epithelial cells from uninfected volunteers and symptomatic patients. J Clin Microbiol 2006;44:2265–7. https://doi.org/10.1128/JCM.02055-05.

[18] European Centre for Disease Prevention and Control. Options for the use of rapid antigen tests for COVID-19 in the EU/EEA and the UK, https://www.ecdc.europa.eu/en/publications-data/options-use-rapid-antigen-tests-covid-19-eueea-and-uk [accessed 22 December 2020].

[19] Larremore DB, Wilder B, Lester E, Shehata S, Burke JM, Hay JA, et al. Test sensitivity is secondary to frequency and turnaround time for COVID-19 screening. Sci Adv 2020;eabd5393. https://doi.org/10.1126/sciadv.abd5393.

[20] Bullard J, Dust K, Funk D, Strong JE, Alexander D, Garnett L, et al. Predicting infectious severe acute respiratory syndrome coronavirus 2 from diagnostic samples. Clin Infect Dis 2020;ciaa638. https://doi.org/10.1093/cid/ciaa638.

[21] Otsuka Pharmaceutical Co., Ltd.. Quick Navi™-COVID 19 Ag Rapid diagnostic test in Japan now enables test sample collection from front of nasal cavity, https://www.otsuka.co.jp/en/company/newsreleases/2020/20201005_1.html [accessed 22 December 2020].

