## Supplementary figures and images for "The evaluation of a newly developed antigen test (QuickNavi™-COVID19 Ag) for SARS-CoV-2: A prospective observational study in Japan"

### Supplementary Figure 1

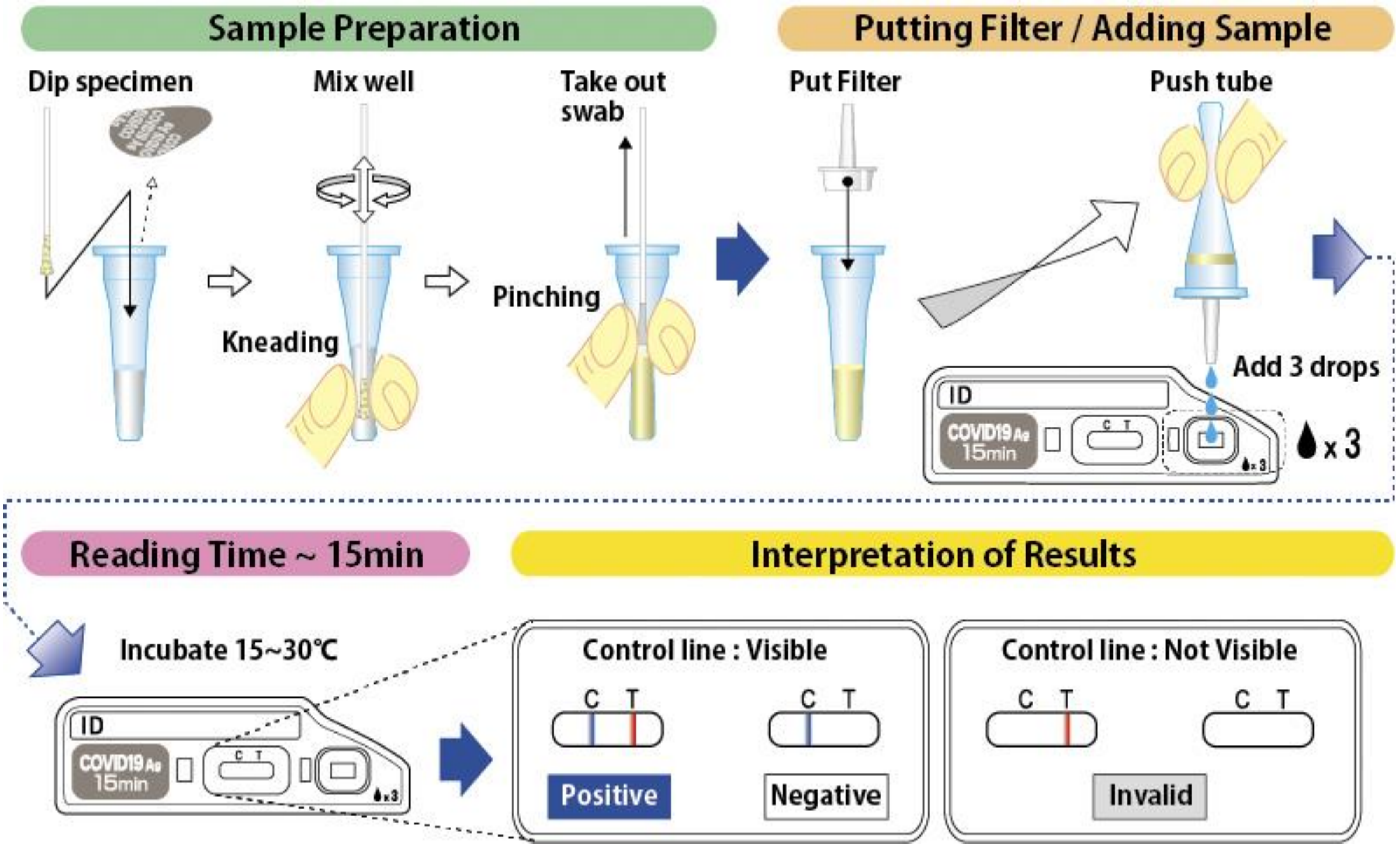


Supplementary Figure 1. Manufacturers' instructions for the QuickNavi™-COVID19 Ag
