## Supplementary Figure 2 for "The evaluation of a newly developed antigen test (QuickNavi™-COVID19 Ag) for SARS-CoV-2: A prospective observational study in Japan"

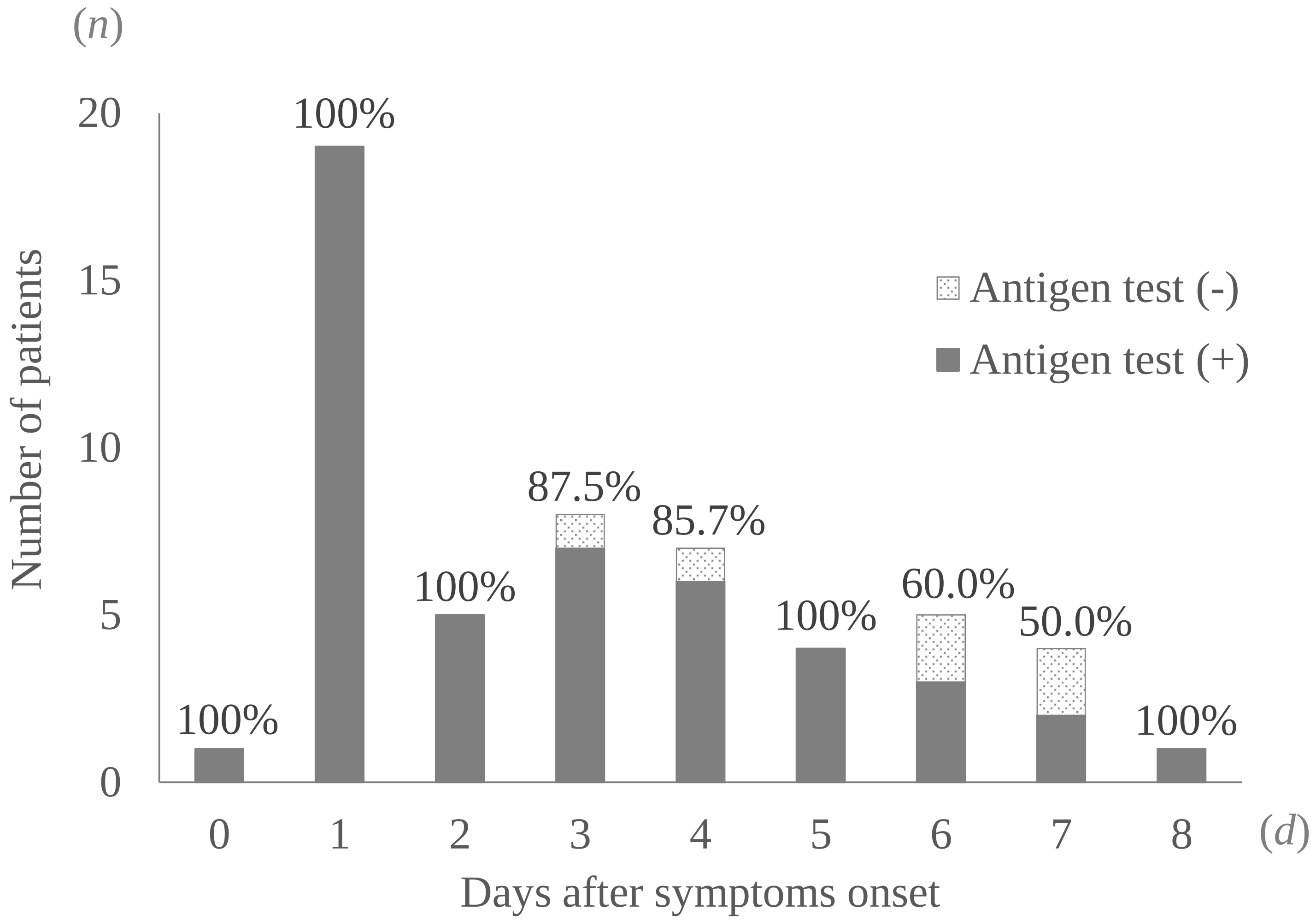


Supplementary Figure 2. Differences in the sensitivity of antigen test stratified by the duration since symptom onset
