## Supplementary Table 1 for "The evaluation of a newly developed antigen test (QuickNavi™-COVID19 Ag) for SARS-CoV-2: A prospective observational study in Japan"

**Supplementary Table 1. Primary facilities and numbers of patients**

|  | Primary facilities | Number of patients |
| --- | --- | --- |
|  | Local health center | 928 |
|  | Clinic A | 159 |
|  | Clinic B | 101 |
|  | Clinic C | 62 |
|  | Clinic D | 60 |
|  | Clinic E | 54 |
|  | Clinic F | 43 |
|  | Clinic G | 42 |
|  | Clinic H | 40 |
|  | Clinic I | 36 |
|  | Clinic J | 32 |
|  | Clinic K | 31 |
|  | Clinic L | 30 |
|  | Clinic M | 30 |
|  | Clinic N | 27 |
|  | Clinic O | 25 |
|  | Clinic P | 24 |
|  | Clinic Q | 21 |
|  | Clinic R | 19 |
|  | Clinic S | 17 |
|  | Clinic T | 16 |
|  | Clinic U | 12 |
|  | Clinic V | 12 |
|  | Clinic W | 12 |
|  | Clinic X | 11 |
|  | Clinic Y | 11 |
|  | Clinic Z | 8 |
|  | Clinic AA | 8 |
|  | Clinic AB | 8 |
|  | Clinic AC | 8 |
|  | Clinic AD | 8 |
|  | Clinic AE | 8 |
|  | Clinic AF | 7 |
|  | Clinic AG | 7 |
|  | Clinic AH | 7 |
|  | Clinic AI | 7 |
|  | Clinic AJ | 6 |
|  | Clinic AK | 6 |
|  | Clinic AL | 6 |
|  | Clinic AM | 6 |
|  | Clinic AN | 6 |
|  | Clinic AO | 6 |
|  | Clinic AP | 6 |
|  | Clinic AQ | 6 |
|  | Clinic AR | 5 |
|  | Clinic AS | 5 |
|  | Clinic AT | 4 |
|  | Clinic AU | 4 |
|  | Clinic AV | 4 |
|  | Clinic AW | 4 |
|  | Clinic AX | 4 |
|  | Clinic AY | 4 |
|  | Clinic AZ | 3 |
|  | Clinic BA | 3 |
|  | Clinic BB | 3 |
|  | Clinic BC | 3 |
|  | Clinic BD | 3 |
|  | Clinic BE | 3 |
|  | Clinic BF | 3 |
|  | Clinic BG | 3 |
|  | Clinic BH | 3 |
|  | Clinic BI | 3 |
|  | Clinic BJ | 2 |
|  | Clinic BK | 2 |
|  | Clinic BL | 2 |
|  | Clinic BM | 2 |
|  | Clinic BN | 2 |
|  | Clinic BO | 2 |
|  | Clinic BP | 2 |
|  | Clinic BQ | 2 |
|  | Clinic BR | 1 |
|  | Clinic BS | 1 |
|  | Clinic BT | 1 |
|  | Clinic BU | 1 |
|  | Clinic BV | 1 |
|  | Clinic BW | 1 |
|  | Clinic BX | 1 |
|  | Clinic BY | 1 |
|  | Clinic BZ | 1 |
|  | Clinic CA | 1 |
|  | Clinic CB | 1 |
|  | Clinic CC | 1 |
|  | Clinic CD | 1 |
|  | Clinic CE | 1 |
|  | Clinic CF | 1 |
|  | Clinic CG | 1 |
|  | Clinic CH | 1 |
|  | Clinic CI | 1 |
|  | Clinic CJ | 1 |
|  | Clinic CK | 1 |
| Total | 90 | 2079 |
